# Abnormal hippocampo-cortical theta-gamma phase-amplitude coupling in Alzheimer’s disease

**DOI:** 10.64898/2026.02.06.26345635

**Authors:** Erfan Baradarantohidi, Srikantan S Nagarajan, Kamalini G Ranasinghe, Stefan Haufe

## Abstract

**INTRODUCTION:** Cognitive decline in Alzheimer’s disease (AD) may arise not only from neuronal loss but also from disrupted temporal coordination across distributed networks. Prior works in healthy individuals has shown memory-related modulation of hippocampal–cortical phase–amplitude coupling (PAC). We hypothesized alteration of resting-state hippocampo-cortical PAC in AD which relates to cognitive impairment.

**METHODS:** Resting-state magnetoencephalography (MEG) data were obtained from 78 AD patients and 70 healthy controls. Source-reconstructed hippocampal and cortical signals were analysed using antisymmetrized bispectral methods to quantify non-linear across-site PAC.

**RESULTS:** AD patients showed significant disruptions of hippocampo-cortical PAC, particularly involving frontal, parahippocampal, and posterior-cingulate cortices. PAC measures were significantly associated with Mini-Mental State Examination scores within the AD group, with higher PAC corresponding to greater cognitive impairment in regions showing increased coupling relative to controls.

**DISCUSSION:** These findings demonstrate altered hippocampo-cortical temporal coordination in AD and suggest PAC as a potential electrophysiological marker of disease-related network dysfunction.

## 1. BACKGROUND

Alzheimer’s disease (AD), the most common neurodegenerative disorder, affects tens of millions of people globally, with prevalence continuing to rise while the efficacy of disease-modifying therapies remains limited.^1^ The cardinal clinical manifestation of AD is a progressive impairment of memory and other cognitive domains, including executive function, visuospatial abilities, and language. These deficits are linked not only to neuronal loss but also to alterations in the coordinated activity of neural networks.^2^

Indeed, traditional spectral and coherence analyses of electrophysiological recordings have provided valuable insights into power alterations across frequency bands in AD.^3–8^ However, recent results suggest that local and non-local nonlinear interactions of electrophysiological activations across brain regions – in particular phase-amplitude coupling (PAC) – may serve as a more sensitive marker of network dysfunction in AD.^9^ PAC quantifies the relationship between the phase of low-frequency oscillations (such as theta or alpha) and the amplitude of high-frequency activity (often in the gamma range). As a form of cross-frequency coupling, PAC can be classified as either within-site or across-site, depending on whether the phase and amplitude signals originate from the same or different brain regions. Across-site PAC is of particular interest because it provides a measure of temporally and spatially coordinated neuronal activity across brain regions.

PAC has been linked to essential cognitive processes in the hippocampus, such as episodic memory and control of working memory.^10,11^ Importantly, among the affected neural circuits in AD, the hippocampus plays a central role, as functions such as memory consolidation critically depend on it and its communication with distributed cortical areas.^12–14^ In AD, the hippocampus is one of the brain regions most impacted in the early phases of the disease – a rapid loss of hippocampal tissue correlates with a functional disconnection from other brain areas.^15–19^ As AD advances, the progressive atrophy of the medial temporal and hippocampal regions becomes evident, serving as key structural indicators in magnetic resonance imaging.^20^ Previous studies have confirmed the feasibility of non-invasive magnetoencephalography (MEG) recording to detect subcortical oscillations.^19,21–24^ However, to date, only one study using animal models of AD has investigated PAC involving the hippocampus.^25^

When using non-invasive measurements such as magnetoencephalography (MEG), conventional methods for measuring PAC (*e.g.* the modulation index)^26^ face challenges in distinguishing genuine across-regional coupling from spurious coupling introduced by signal leakage from a source containing within-site coupling. Consequently, a bispectral approach was applied to estimate PAC in the present study, which circumvents this limitation of conventional methods by leveraging higher-order statistics. Bispectral methods reduce computational complexity and are not dependent on parameters like the filter width. In addition, the anti-symmetrized bispectrum, looking at the part of the bispectrum that is antisymmetric to the permutation of two channels, offers improved robustness in detecting genuine across-regional interactions.^27,28^

This study investigates whether Alzheimer’s disease (AD) is associated with impaired phase-amplitude coupling (PAC) between the hippocampus and cortical regions during resting-state activity. Based on evidence suggesting early disruption of hippocampal oscillatory dynamics in AD, we test the overall hypothesis that hippocampo-cortical PAC is selectively altered in individuals with AD compared to cognitively healthy controls. To evaluate this hypothesis, we address the following research questions:

1. Does AD affect the communication between hippocampal and cortical oscillations as measured by robust across-site phase-amplitude coupling during the resting state?
2. If so, what cortical regions show significant hippocampal PAC alterations in AD, and how do these relate to the known regional vulnerabilities associated with the disease’s clinical features?
3. Is impaired PAC associated with cognitive decline as indexed by clinical scores (*e.g.* MMSE)?

## 2. METHODS

The present study is based on data acquired at the Alzheimer’s Disease Research Center at the University of California, San Francisco (UCSF).^29^

### 2.1 Dataset

The following information is adapted from Kudo *et al.*^29^. The original study involved 78 individuals (50 female; 28 male) diagnosed with AD according to the National Institute of Aging–Alzheimer’s Association guidelines,^30–32^ alongside 70 older adults (41 female; 29 male) with no cognitive impairment. Subjects were recruited from research cohorts at the UCSF Alzheimer’s Disease Research Center. A multidisciplinary team confirmed the AD diagnoses through consensus. Both participant groups completed the Mini-Mental State Examination (MMSE) and a structured interview with caregivers to evaluate the Clinical Dementia Rating (CDR) scale. Control participants scored zero on the CDR, reflecting no cognitive deficits, whereas AD patients scored between 0.5 and 2. Additional information is available in Kudo *et al.*^29^.

Brain structural imaging was conducted using a standardized MRI protocol on 3T Siemens scanners (MAGNETOM Prisma or 3T TIM Trio) at the UCSF Neuroscience Imaging Center. Scans occurred, on average, 1.05 years (range: -6.91 to 0.78) and 0.29 years (range: -2.13 to 1.29) before or after MEG assessments for controls and patients, respectively.

Between 10 and 60 minutes of resting-state MEG data were collected per participant at the UCSF Biomagnetic Imaging Laboratory using a 275-channel CTF Omega 2000 system (CTF MEG International Services LP, Coquitlam, British Columbia, Canada). Participants’ data were recorded in a supine position with eyes closed (sampling rate *f*_*s*_ = 600 Hz). Three fiducial coils were placed at the nasion and bilateral preauricular points to track head position relative to the sensor array, later co-registered with individual MRI scans. Efforts were made to limit head movement during sessions to under 0.5 cm. A 1-minute continuous segment with minimal artifacts – characterized by minimal excessive scatter at signal amplitude – was chosen per subject from the continuous recordings.

The 1-minute MEG sensor data were filtered digitally with a band-pass filter in the range of 0.5-55 Hz. The power spectral density (PSD) was calculated for each sensor, and visual inspections confirmed the presence of artifacts. Noisy channels were excluded from further analysis within each participant’s dataset. A preconditioned independent component analysis (ICA)^33^ was then used to isolate and eliminate cardiac artifacts. In each recording, one or two ICA components displaying clear cardiac waveforms with approximately 1 Hz rhythms were detected and removed following visual confirmation.

Further preprocessing steps were employed using the FieldTrip toolbox.^34^ The data were down sampled to 200 Hz and split into epochs of 2 seconds length with no overlap. Outlier channels and epochs – defined as exceeding ±3 standard deviations from the median – were identified, with outlier channels subsequently interpolated using the average of their neighbouring channels. Outlier epochs were rejected.

### 2.2 Forward Model

It is assumed that most, but not all MEG/EEG signals are generated by the postsynaptic activity of ensembles of cortical pyramidal neurons of the cerebral cortex. The reason lies in the morphology and mass effect of these cells, which have elongated shapes and are grouped in large assemblies oriented in a similar manner approximately normal to the cortex. The first step to source estimation (mapping sensor-level recordings to brain activity and estimating neural currents generated by the postsynaptic activities) involves modelling the electromagnetic properties of the head (electrical volume conductor models) and the sensor array, which builds up the forward model, resulting in the leadfield or gain matrix. It defines the linear relationship between neural current sources and measurements at the sensor level. The pipeline used in this study involves the creation of realistic individual head models based on the co-registered MRI, placing dipolar primary neuronal current sources equidistantly halfway between the inner and outer cortical surfaces and on the hippocampus surface, importing the sensor channel’s location description and computing the leadfield matrices using the Boundary Element Method (BEM). We start by describing the construction of individual participants’ head models using available structural brain images.

#### 2.2.1 Individual Head Modelling

Individual T1 MRI structural brain images were processed by the *recon-all* pipeline of the structural MRI analysis software FreeSurfer^35^ to obtain high-resolution 3D surface meshes of the outer cortical surface (the interface between cortical grey matter, GM, and cerebrospinal fluid, CSF) as well as the inner cortical surface (the interface between GM and white matter, WM). In addition, an individual parcellation of the cortex to 68 regions-of-interest (ROI) according to the Desikan-Killiany anatomical atlas (DK)^36^ and an automatic subcortical segmentation of brain volume (Aseg)^37^ were obtained.

Two subjects had MRI images with stripe artifacts that could not be corrected or processed by FreeSurfer, leading to their exclusion from the analysis.

#### 2.2.2 Mixed Source Modelling

The source space in this study consists of the surface of the cortex and the subcortical structures; hence, we speak of a mixed source model. The cortical source space was modelled by placing 5000 current dipoles at cortical locations halfway between the inner and outer cortical surfaces with normal orientation to the outer surface. Deep structures such as the amygdala, the thalamus, and the hippocampus were part of the Aseg atlas extracted by Freesurfer.^35^ Supplementary Fig. 1 is a visualization of the extracted subcortical structure volumes. Based on recommendations from Attal *et al.*^21^, hippocampal sources were positioned on the hippocampal surface with a normal orientation, similar to the approach used for cortical sources. This procedure was implemented in MATLAB (MathWorks Inc., Natick, USA) using low-level functions of the Brainstorm toolbox.^38^

The overall number of subcortical sources was set to a quarter of the cortical sources. Although only the hippocampi were used in source-level analysis, other subcortical structures such as the brainstem, cerebellum, accumbens, amygdala, caudate, putamen, and thalamus were also modelled. The exact number of hippocampal sources per subjects varied due to anatomical differences.

#### 2.2.3 Leadfield computation

The forward problem solving or leadfield computation in MEG amounts to calculating the theoretical magnetic fields generated by current sources in the brain. Maxwell’s equations govern electromagnetic propagation, coupling the electrical and magnetic fields. This coupling simplifies when the relevant frequencies are low enough for the quasistatic regime to hold. The OpenMEEG software package^39^ implementing a symmetric Boundary Element Method (BEM) was used to compute the forward magnetic fields in that regime.^40^ The leadfield defines the contribution of each source to every sensor, and for each source, it is represented as a 3-by-number of sensors vector, corresponding to the three spatial directions.

Using the Brainstorm toolbox,^38^ three concentric triangularised surfaces (referred to as 3 BEM layers/shells) defining separate compartments for brain, skull, and scalp tissue, were derived from the individual structural MRI. MR images were co-registered with CTF axial gradiometers sensor locations and normalized to the Montreal Neurological Institute (MNI) coordinate system.^41^ Piecewise constant electrical conductivity fields of the nested tissues (defined by the three concentric surfaces) *σ*_brain_ = 0.33*S*/*m*, *σ*_skull_ = 0.041*S*/*m*, and *σ*_scalp_ = 0.33*S*/*m*, respectively, were assumed for the brain, skull, and scalp compartments.

The extracted surface and sensor locations with overlaid MRI data were visualized and manually inspected for artifacts and errors such as improper head surface shape and unsuccessful recognition of inner/outer surfaces; instances for one subject are shown in Supplementary Fig. 2.

In case of an overlap between any of the outer cortex surface and the 3 BEM layers, small adjustments to the vertices of the surface envelope were applied to force the outer surface inside the inner one using the Brainstorm toolbox. Despite this procedure, the extracted surfaces for six subjects could not be fixed; therefore, only the inner skull layer was used for those subjects.

### 2.3 Inverse Modelling

To reconstruct source activity, the linearly-constrained minimum variance (LCMV) beamforming algorithm was applied to the MEG sensor measurements using individual leadfields.^43^ To prevent rank deficiency, the sensor space covariance matrix ***C*** ∈ ℝ^*N*×*N*^ was regularized by adding 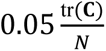 to each diagonal element before the application of LCMV. Since MEG is insensitive to purely radial sources, keeping radial component would introduce redundancy, numerical instability, and computational inefficiency in source reconstruction. The radial dimension of the leadfields was, therefore, removed by applying Singular Value Decomposition (SVD) to each source’s leadfield and retaining only the first two singular components, which capture the tangential directions. These components are not restricted to the x-, y-, or z-axes but span a free tangential plane at each source location, discarding the local radial component. The beamforming procedure was implemented using custom scripts in MATLAB.

#### 2.3.1 Regional Signal Aggregation and Power

The mixed source space was divided into 68 cortical ROIs according to the Desikan-Killiany anatomical atlas^36^ and the left and right hippocampi in the Aseg atlas.^35^ Since every ROI contains multiple sources, the estimated activation time courses of all estimated sources within each region’s x-, y- and z-components were summarized using principal component (PC) analysis. Only the first three PCs of each region,^44^ sorted by variance explained, were retained for further analysis. In addition to the regional time series, the cross-spectrum for all sources in that region was computed using 2-second-long epochs (frequency resolution, 0.5 Hz) multiplied with a Hanning window before applying the fast Fourier transform. The regional power spectral density at a given frequency was then extracted as the sum of the diagonal elements of the source × source cross-spectral matrix divided by the number of sources.

### 2.4 Bispectral analysis and PAC

The third-order statistical moment of the Fourier transform is called the cross-bispectrum. The general cross-bispectrum *B* between channels *k*, *m*, *n* and at low frequency *f*_1_ and high frequency *f*_2_ can be written as

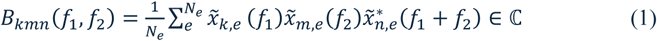

where *x̃*_*k*,*e*_(*f*) represents the *e*-th segment of the Fourier-transformed data of channel *k* at frequency *f*, and where ∗ denotes complex conjugation.

Close relation between conventional PAC estimates^26,45^ and the bispectrum have been pointed out by several studies.^46–48^ In short, the cross-bispectrum (1) is a suitable measure of coupling of the phase of the signal at channel *k* and low frequency *f*_1_ and the amplitude of the signal at channel *m* and high frequency *f*_2_,

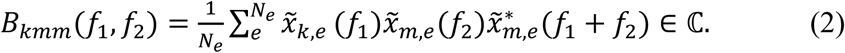

Similar to coherence, which is the normalized version of a cross-spectrum,^49^ bicoherence can be defined as a normalized version of the cross-bispectrum,

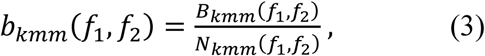

where the normalization factor, introduced by Shahbazi *et al.*^50^, is given by

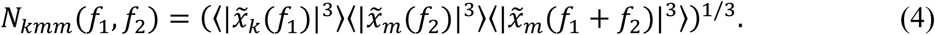

The normalization factor in (4) is not affected by coupling between signals and bicoherence derived using this factor in (3) has been shown to be bounded by zero and one.^50^

According to Zandvoort and Nolte^47^, the coupling between the phase of a slow oscillation at *f*_*slow*_ and the amplitude of a fast oscillation at *f*_*fast*_ corresponds to two bispectral estimates 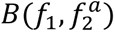 and 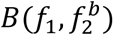, where *f*_1_ = *f*_*slow*_, 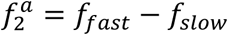 and 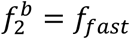. When PAC is present in the signal, two additional peaks of spectral power (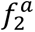 and *f*_*fast*_ + *f*_*slow*_) around the peak of the fast oscillation, *f*_*fast*_, are present. Consequently, the average of 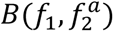 and 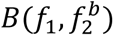 is then a better estimate of PAC.

The use of bicoherence to examine the interaction between regions is confounded by the source leakage, source leakage refers to the mixing of signals from different brain regions, causing a single source to be inaccurately represented as multiple or spreading its activity to neighbouring regions. To overcome this problem the Bispectral anti-symmetrization was introduced by Chella *et al.*^27^. They demonstrated that anti-symmetrized bispectra vanish for mixtures of independent sources. If slow and fast oscillations originate from separate sites, suggesting a true interaction across sites, the slow oscillation would be confined to one site, and the fast oscillation would be observed at the other. In contrast, if the PAC interaction originates from a single source that spreads to two sites, both the slow and fast oscillations would be present at each site. By leveraging this distinction, genuine interactions can be identified by isolating the anti-symmetric component and removing the symmetric part of the interaction.

Consequently, Pellegrini *et al.*^28^ proposed to use anti-symmetrized bispectra to estimate genuine across-site PAC (ASB-PAC) and to distinguish it from spurious across-site PAC that could potentially originate from a single source. In their simulation study, Pellegrini *et al.*^28^ validated ASB-PAC by comparing it to bispectral estimates and conventional MI-based PAC estimates and observed that ASB-PAC achieves the highest performance in detecting genuine across-site PAC interactions while detecting the fewest spurious (within-site) interactions. The anti-symmetrization is performed by subtracting the symmetric part of the cross-bispectrum, *B*_*mkm*_from the cross-bispectrum in (2).

Taken together, the approach used in this study to estimate the across-site PAC between channels *k* and *m* at frequencies *f*_*slow*_ and *f*_*fast*_, respectively, is to compute the average of the absolute values of anti-symmetrized bicoherence at two frequency settings, *a* and *b*:

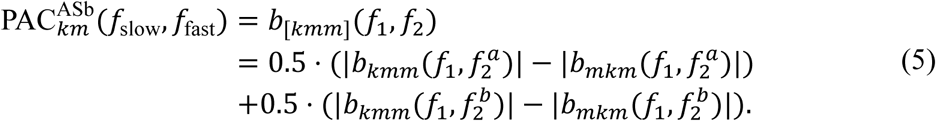

The bispectrum was computed using the available 60-second source data, with one 60-second epoch divided into 1-second segments (providing a frequency resolution of 1 Hz) that is shifted in increments of 0.2 seconds, resulting in a 0.8-second overlap between consecutive segments. Each segment was multiplied by a Hanning window before applying the fast Fourier transform.

As mentioned earlier, the first three principal components of the sources within each of the 68 ROIs in the DK atlas and each seed region (both hippocampi) were extracted; therefore, in every ROI-seed pair connection, PAC in 9 different channel combinations can be computed. For both seed regions and each ROI, the phase of the signals in the theta and alpha (4-12 Hz) frequency ranges, and the amplitude of the signals in the low gamma (30-55 Hz) range were analysed. Note that the sensor data are low-pass filtered at 55 Hz, and due to the third term in (2) that is the Fourier coefficient of the data in frequency *f*_1_ + *f*_2_, not all theta-alpha versus gamma PAC values are valid (as *f*_1_ + *f*_2_ might be higher than the 55 Hz).

### 2.5 Statistical Analysis

#### 2.5.1 Linear Regression

To quantify regional and band-specific power alteration, a linear regression model (6) was fitted independently for each of the 68 ROIs in the DK anatomical atlas in the delta-theta (2 to 7 Hz), alpha (8 to 12 Hz), and beta (15 to 29 Hz) frequency bands. The response variable of the models was the relative band power of subjects within both groups. The predictor of interest was group membership while we adjusted also for the effect of age.

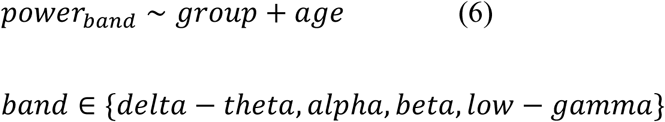

In every frequency band, 68 models corresponding to 68 ROIs were fitted. A significant *t-*statistic (defined by a *P-*value < 0.01) for the coefficient estimate of *group* quantifies the regional changes of the relative power in the AD group from the control group. To account for the multiple comparison problem, the *P-*values were adjusted by the false discovery rate (FDR) controlling procedure introduced by Benjamini and Hochberg ^51^.

#### 2.5.2 Across-site PAC Group Differences

To balance statistical rigor and exploratory flexibility, both linear mixed-effects (LME) model and cluster-based permutation test (CBPT) were employed to examine group differences in hippocampo-cortical PAC at the regional level. The subsequent section outlines the LME modelling approach, whereas the section following after describes a CBPT, a nonparametric statistical test suggested for analysing EEG- and MEG-data providing a straightforward way to solve the multiple comparisons problem (MCP). The LME model and the CBPT were applied to the raw PAC values derived from each ROI’s first principal component. For each ROI, due to the band-pass filter’s 55 Hz upper limit, only 147 of the 234 (9 × 26) theta–alpha–gamma frequency pairs were valid. The multiple comparison problem for the LME model is addressed by FDR correction using a desired false discovery rate of *q* = 0.05. Since our primary interest lies in the effect of AD, positive effects sizes indicate an increase relative to controls and vice versa.

#### 2.5.3 Linear Mixed-Effects Model

A linear mixed-effects model was designed to statistically test group differences for each region combination. Subject-level and frequency-level sources of variability were modelled using random intercepts grouped by each variable. The variable of interest, group membership, was modelled as a fixed effect. The Wilkinson notation^52^ of the LME model is

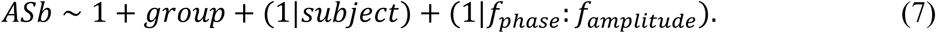

Here, *ASb* is the continuous response variable (PAC as derived from anti-symmetrized bicoherence), while all predictor variables are categorical. The output of model (7) for the fixed-effect term includes one intercept estimate and one coefficient estimate for the group membership.

For each ROI, the coefficient estimate of the *group* was statistically tested using a *t-*test. To account for multiple comparisons, the *P-*values associated with the group coefficient estimates from each ROI were adjusted using the false discovery rate (FDR) method.^51^ Group-related differences correspond to mean differences across all frequency pairs, while deviations from this mean are captured by the random effect terms (1|*f*_*phase*_: *f*_*amplitute*_). A corrected alpha level of *P* < 0.05 was used to define a significant difference in across-site PAC between the AD and control groups.

#### 2.5.4 Cluster-Based Permutation Test

Following the approach of Prabhu *et al.* ^9^, a cluster-based permutation test^53,54^ was also applied to assess group differences in across-site PAC. For each frequency-frequency pair, PAC contrast was estimated using an independent two-sample *t-*test with unequal variance assumption.^55^ Then, all frequency-frequency pairs whose *t-*values are larger than ±1.6 (below a cluster alpha threshold of 5 %) are selected. Selected frequency-frequency pairs are assigned to clusters based on connectedness in both phase and amplitude frequencies. Cluster-level statistics are calculated by taking the sum of the *t-*values within every cluster. A null distribution of contrast statistics was calculated based on 1024 permutations (or repetitions) in which group labels are randomly reassigned across PAC values for each region. The permutation *P-*value was found by contrasting the cluster statistic with the null distribution. When the total of the *t-*values was larger than the 95th percentile (corresponding to a cluster *P-*value < 0.05) of the permuted distribution, the observed clusters were regarded as statistically significant.

#### 2.5.5 Correlation

We investigated the correlation between regional across-site PAC values and cognitive function as measured by MMSE scores in the AD group. For each hippocampus, we limited our analysis to the ROIs that showed significant alteration in hippocampo-cortical PAC in the cluster-based permutation test and averaged PAC values across the significant cluster(s) of the frequency-by-frequency tiles. The correlation between averaged PAC values and MMSE score were assessed using Pearson’s linear correlation coefficient (*r*).

### 2.6 Code and Data Availability

The dataset used in this study, including de-identified and preprocessed MEG recordings and structural brain images (MRI), is publicly available at OSF (https://doi.org/10.17605/OSF.IO/PD4H9). All analysis scripts supporting the results of this study are openly accessible at our GitHub repository (https://github.com/braindatalab/AD_PAC).

## 3. RESULTS

### 3.1 Spectral Power

The power spectral density averaged over all cortical regions and across both hippocampi in the range from 1 to 55 Hz are shown in Fig 1. Before averaging over all regions, the PSD for each group was normalized by dividing the subjects’ power in each region by the mean power within the 0 to 55 HZ frequency range in that region.

**Figure 1.**
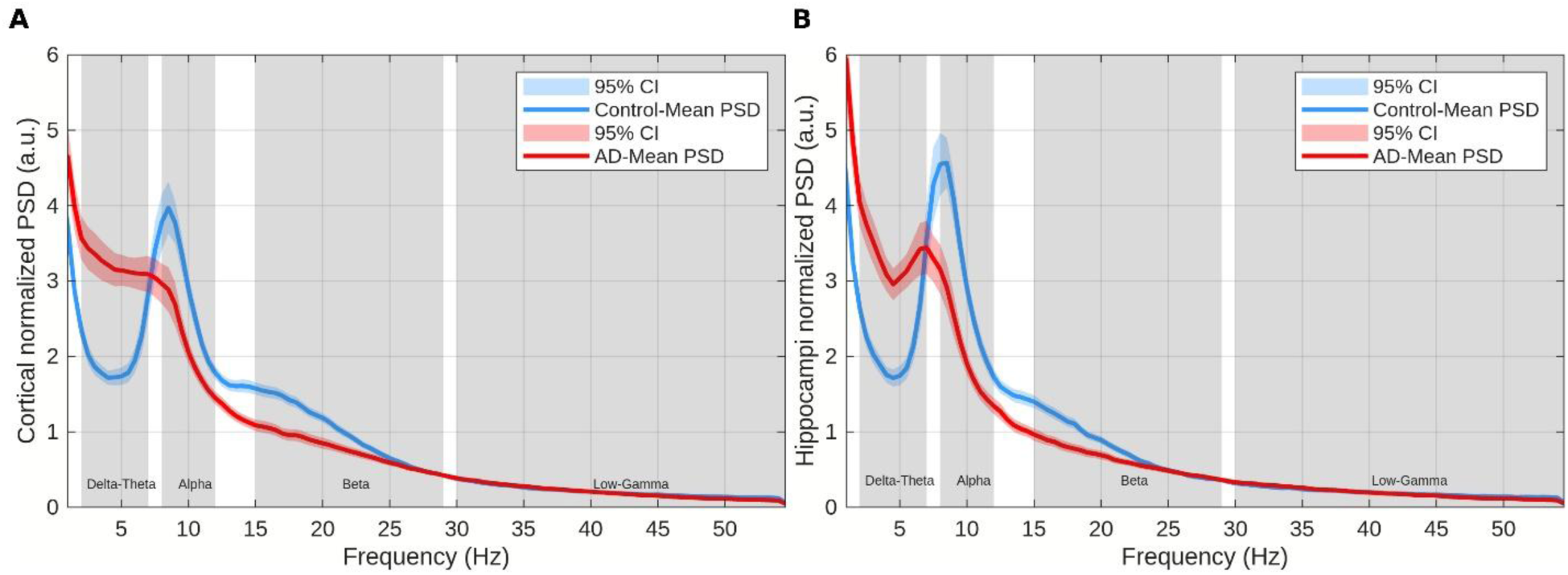
Normalized power spectral densities (PSDs) for the Alzheimer’s disease (AD) and control groups. **(A) Cortical** power spectral densities. **(B) Hippocampal** power spectral densities. The solid red and blue lines represent the group mean PSD, and the red and blue shaded areas are the 95 % confidence intervals for the AD and control groups, respectively.

Abnormal power alterations in AD compared to age-matched cognitively unimpaired controls have been reported in many studies.^5–8,19,29^ Here we replicate these results. The observed spectral changes of relative cortical power are band-specific; in particular, we observe an elevation of the delta-theta (2 to 7 Hz) band power and a decrease in alpha (8 to 12 Hz) and beta (15 to 29 Hz) band power. As also described by Kudo *et al.*^29^, alpha peak frequencies of AD patients are generally lower (*i.e.* alpha slowing) compared to controls. The extent of alpha slowing varies by brain region and AD severity. Averaging over PSD curves with different alpha peaks leads to the absence of a common alpha peak in the average PSD for patients with AD (see Fig. 1).

Power alterations observed in hippocampal source estimates are similar to those observed for cortical power; this is in agreement with the resting-state MEG study of Luppi *et al.*^19^, where relative hippocampal power differences in AD, subjective cognitive decline (SCD), and mild cognitive impairment (MCI) were consistent with differences in cortical power. We also observe smaller variance of the hippocampi relative power in the delta-theta range compared to the cortical relative power, confirming the better AD classification performance of hippocampal compared to cortical theta power reported by Luppi *et al.*^19^

Cortical topographic maps of the FDR-corrected significant (*P* < 0.01) *t-*statistic signs multiplied by *log*(*P*) of the *group* in the (6) are shown in Fig. 2. The observed regional group differences of power confirm previous results reporting that the relative power in the delta-theta band is increased across the whole brain, and that alpha band power is decreased in temporal regions.^8,29^ A global beta power decrease is moreover observed. There were no significant regional changes in the low-gamma band.

**Figure 2.**
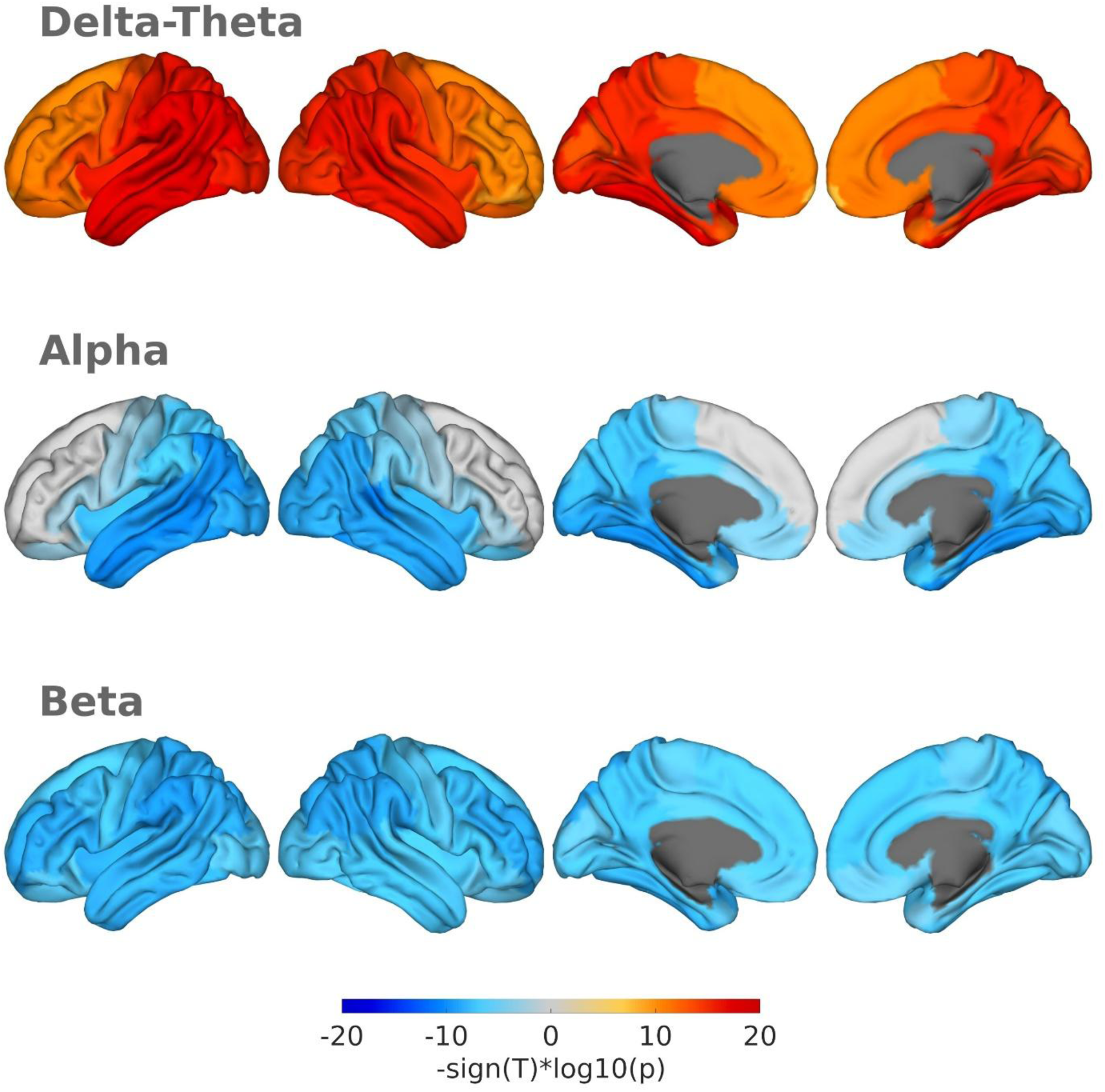
Regions with significant. (*P* < 0.01, FDR corrected) **relative power group differences** (AD vs. Control) **in canonical bands defined as delta-theta** (2 to 7 Hz), **alpha** (8 to 12 Hz), **and beta** (15 to 29 Hz). The colour scale represents the signed statistic, defined as −*sign*(*T*) × *log*₁₀(*p*) with warm colors (red) denoting increased band power in AD relative to controls, and cool colours (blue) denoting reduced band power in AD.

### 3.2 Hippocampo-Cortical PAC

In this section, the results of the main analysis of this study are presented, namely, the effect of AD on theta-alpha vs gamma PAC between hippocampus and cortex.

Parametric and non-parametric statistical models as presented in the Materials and Methods section were applied to the across-site PAC values estimated through anti-symmetrized bicoherence (5), testing group differences at each ROI independently. As mentioned in the Statistical Analysis section, each model was applied for four cases: (1,2) left and right hippocampus as the amplitude provider region (gamma frequencies), (3,4) left and right hippocampus as the phase provider region (theta-alpha frequencies). Results derived from PAC values averaged across left and right hippocampi and a more stringent significance threshold (cluster *P-*value < 0.01) are presented in Supplementary Fig. 3.

Fig. 3 shows the results of the CBPT for the case of hippocampal gamma oscillation being modulated by the cortical theta-alpha oscillation. In contrast to the permutation test, the parametric statistical test based on the LME described in (7) did not detect any significant disruption of hippocampo-cortical interaction in the hippocampal gamma setting. Moreover, the CBPT results on the mean PAC values of the left and right hippocampi did not reveal any group contrast in the hippocampal gamma setting.

**Figure 3.**
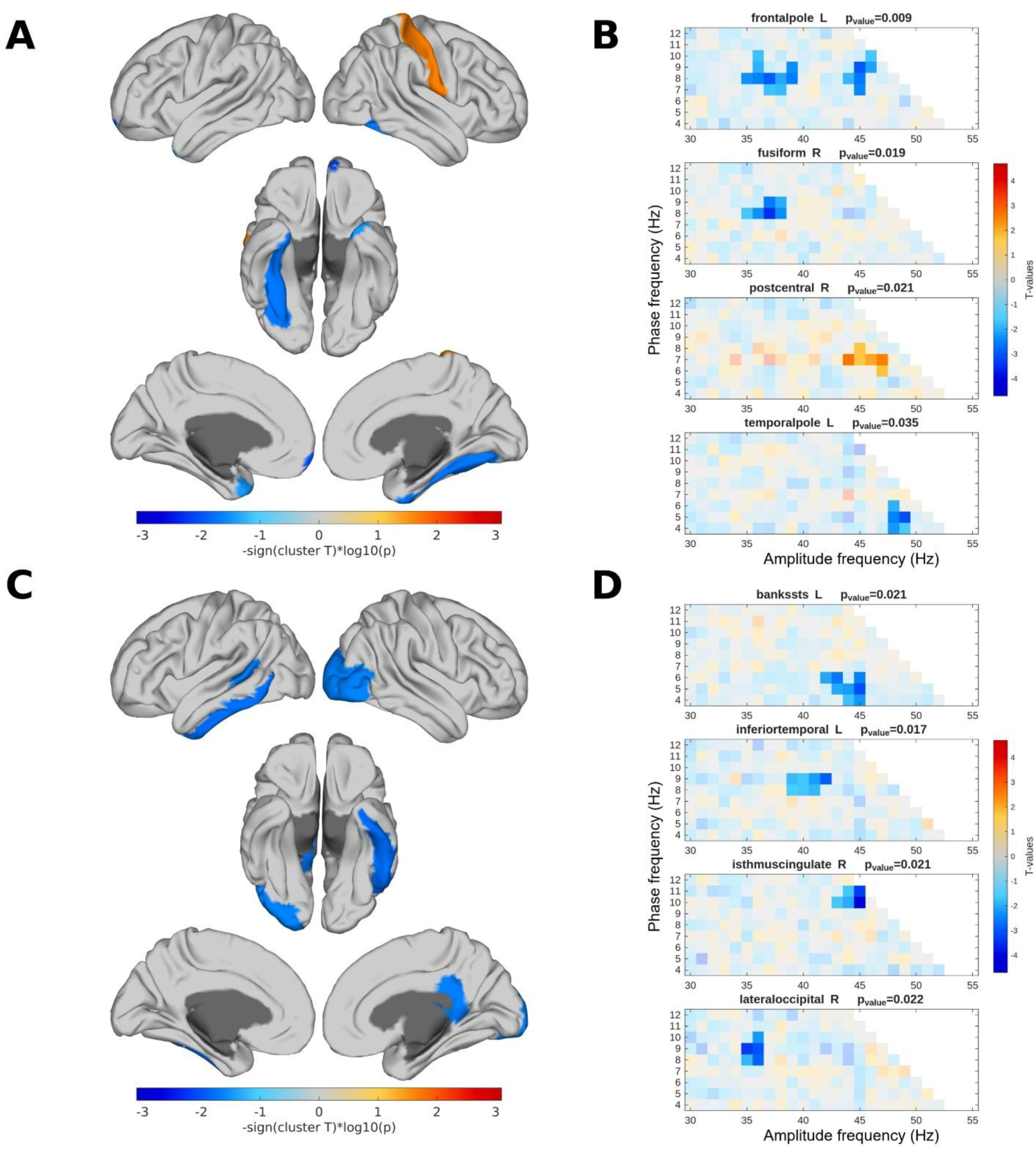
Group differences in across-site phase–amplitude coupling (PAC) with the hippocampus as the amplitude provider region. **(A) Left hippocampus. (C) Right hippocampus.** Cortical regions shown in colour indicate significant clusters (*P* < 0.05) AD vs. control contrast. The colour scale represents the signed cluster statistic, defined as−*sign*(cluster T) × log₁₀(p) with warm colors (red) denoting stronger PAC in AD relative to controls, and cool colors (blue) denoting reduced PAC in AD. **Clusters with significant *t-*statistics observed for different cortical regions in the frequency-by-frequency plane.** In case of multiple significant clusters were detected within a region, the lowest *P-*value is reported. Significant clusters *(P* < 0.05) are shown in full opacity. Clusters are formed with frequency-frequency pairs whose *t-*values are larger (smaller) than ±1.6 (below a cluster alpha threshold of 5 %). **(B) Left hippocampus. (D) Right hippocampus.**

This is different for the case of the hippocampal theta oscillation modulating cortical gamma; both statistical models identified similar – if not the same – regions as being affected in AD (See Fig. 4 and Supplementary Fig. 4). Note that the study of Wang *et al.*^10^ investigating intracranial hippocampal across-site PAC also considered only the case where hippocampal theta oscillation modulates cortical gamma. Furthermore, the fact that parametric and non-parametric tests as well as the both hippocampi averaged PAC values result in the same effects and regions demonstrates the low probability of obtaining the across-site PAC differences for hippocampus as the phase provider under the null hypothesis that AD has no effects on those regions, which is not the evident in our observations for the case of hippocampus as the amplitude provider signal.

**Figure 4.**
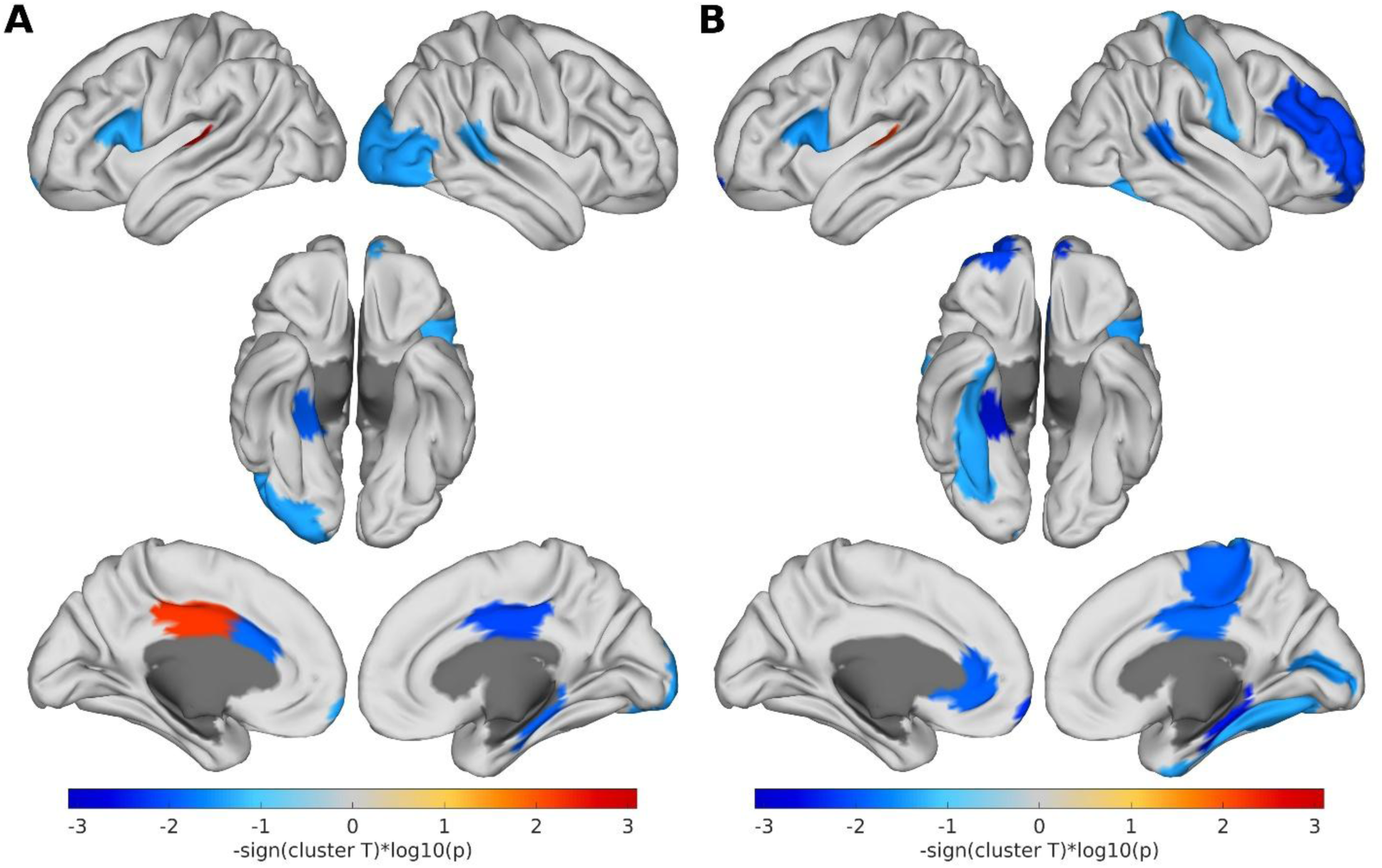
Group difference in across-site PAC, with the hippocampus as the phase provider region. **(A) Left hippocampus. (B) Right hippocampus.** Highlighted regions present a significant cluster (*P* < 0.05) of AD vs. control contrast. The colour scale represents the signed cluster statistic, defined as −*sign*(cluster T) × log₁₀(p) with warm colors (red) denoting stronger PAC in AD relative to controls, and cool colors (blue) denoting reduced PAC in AD.

**Figure 5.**
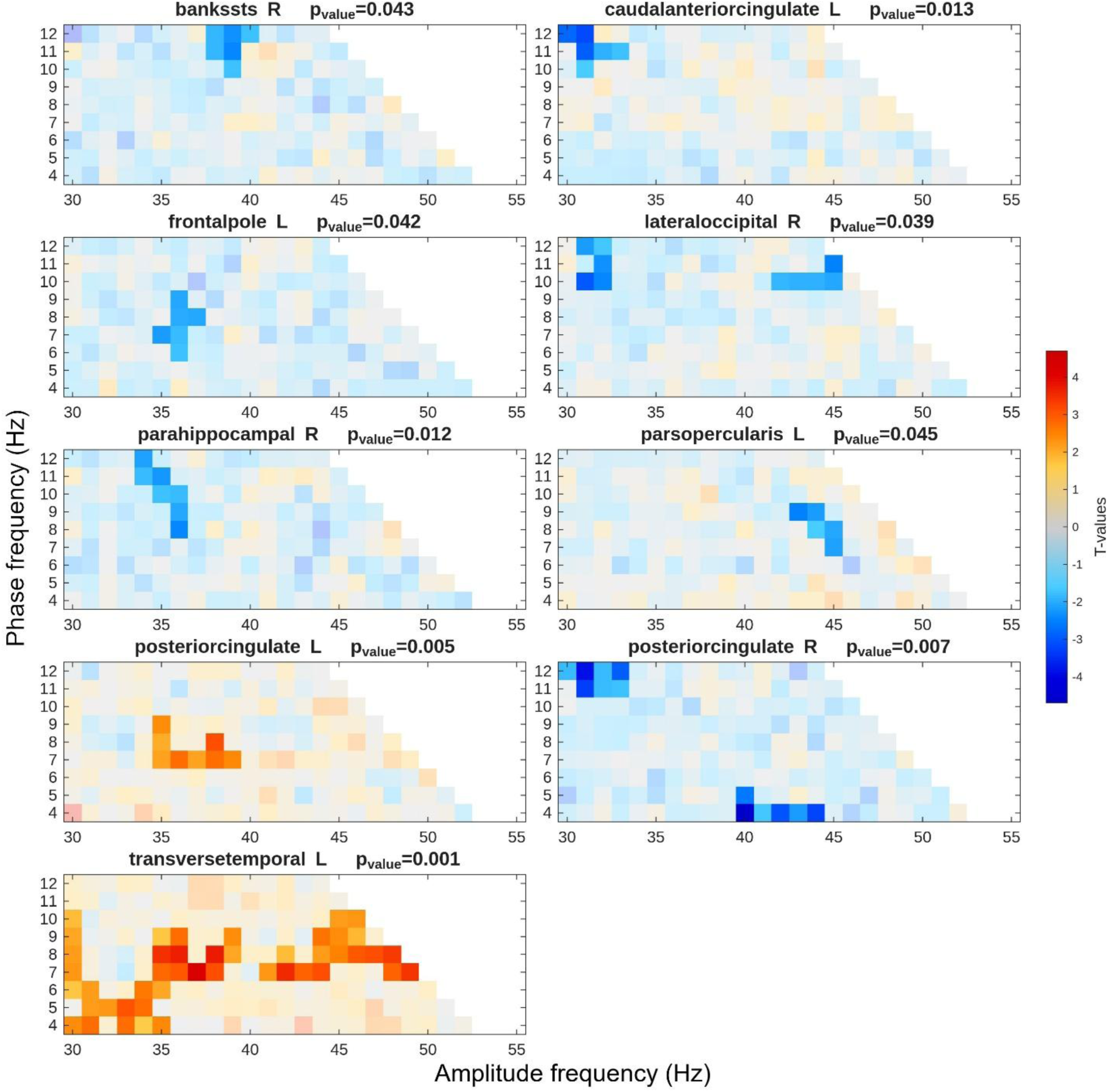
Clusters with significant *t-*statistics observed for different cortical regions in the frequency-by-frequency plane. The **left hippocampus** is taken as the **phase** provider signal in the across-site PAC. In case of multiple significant clusters, the lowest *P-*value is presented. Significant clusters (*P* < 0.05) are shown in full opacity. Clusters are formed with frequency-frequency pairs whose t-values are larger (smaller) than ±1.6 (below a cluster alpha threshold of 5 %).

Furthermore, in the case of hippocampus as the phase provider and considering the left hippocampus, the significant regions shared in CBPT (Fig. 4) and LME (Supplementary Fig. 4) models include the banks of the superior temporal sulcus (right), the frontal pole (left), the lateral occipital (right, two clusters), the parahippocampal (right), the posterior cingulate (right, two clusters), and the transverse temporal (left, two clusters); and for the right hippocampus, the identified regions are the frontal pole (left, four clusters), the fusiform (right), the parahippocampal (right, two clusters), the posterior cingulate (right, two clusters), the rostral middle frontal (right), and the transverse temporal (left, three clusters).

In summary, while cluster-based permutation tests identified significant differences in hippocampal gamma modulation, the LME model did not detect corresponding effects. However, for the case where hippocampal theta-alpha oscillations modulate cortical gamma, both statistical approaches consistently highlighted overlapping regions affected by AD. Notably, significant decreases were observed in regions within the prefrontal, posterior cingulate, and parahippocampal cortices, while increased coupling was detected in the transverse temporal gyrus. These findings highlight significant AD-related differences in hippocampo-cortical interactions, though the underlying mechanisms and functional implications remain to be fully established.

### 3.3 Clinical Score

In each set of ROIs showing alterations in hippocampo-cortical PAC in AD, we found one ROI in which PAC values are correlated with MMSE scores. For the left hippocampus as the phase provider we find a significant Pearson correlations with the average PAC values in the right lateral occipital cortex (*r* = 0.268, *P* = 0.019) while for the right hippocampus as the phase provider we find a significant correlation with the left transverse temporal cortex (*r* = −0.250, *P* = 0.030).

The direction of these effects aligns with differences in hippocampo-cortical PAC between AD patients and controls. In the cortical region (right lateral occipital cortex) exhibiting lower hippocampo-cortical PAC in AD, a positive correlation with MMSE scores was observed—indicating that greater cognitive impairment (*i.e.* lower MMSE scores) was associated with lower hippocampo-cortical PAC values. Conversely, in the region (left transverse temporal cortex) which the AD group showed increased PAC compared to controls, a negative correlation with MMSE scores emerged, suggesting that greater cognitive impairment corresponds to higher PAC values (See Fig. 7).

**Figure 6.**
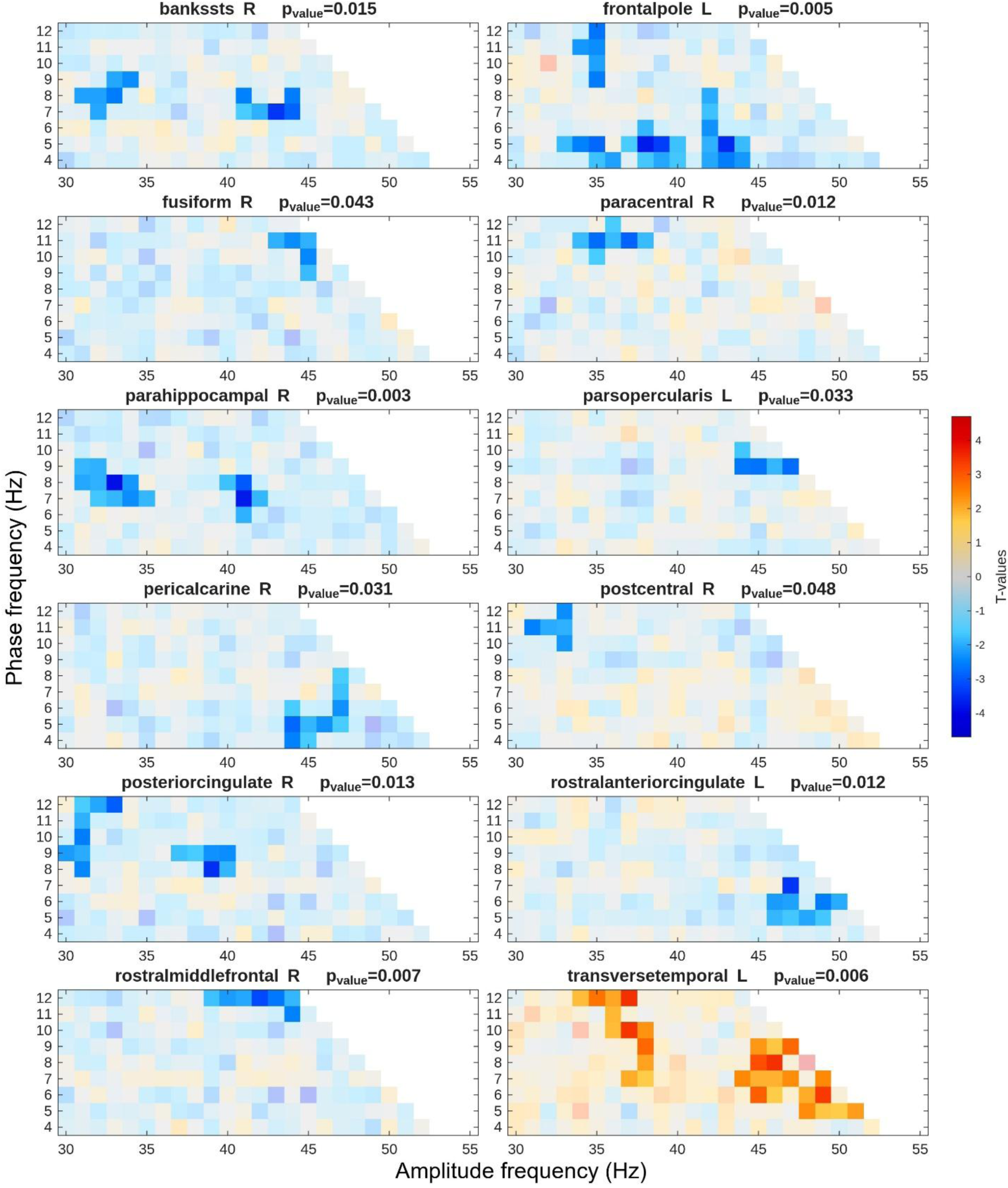
Clusters with significant *t-*statistics observed for different cortical regions in the frequency-by-frequency plane. The **right hippocampus** is taken as the **phase** provider signal. In case of multiple significant clusters, the lowest *P-*value is presented. Significant clusters (*P* < 0.05) are shown in full opacity. Clusters are formed with frequency-frequency pairs whose t-values are larger (smaller) than ±1.6 (below a cluster alpha threshold of 5 %).

**Figure 7.**
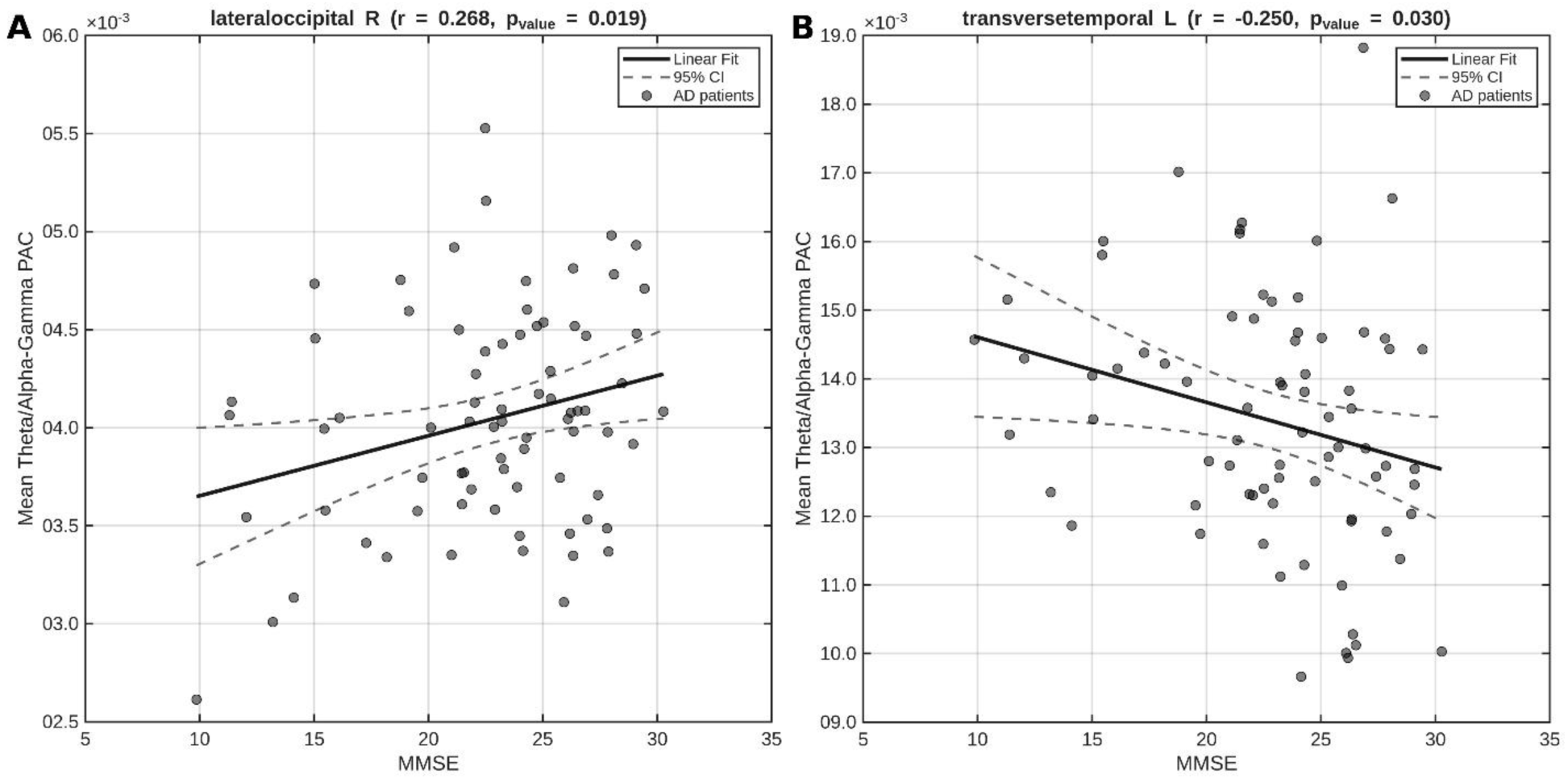
PAC and MMSE scores in AD group. For each hippocampus as **phase** provider signal, one ROI showed significant correlation of the PAC values averaged across the cluster(s) and MMSE scores. **(A)** Positive correlation (*r* = 0.268, *P* = 0.019) in **right lateral occipital cortex** that showed reduction of PAC with **left** hippocampus in AD group. **(B)** Negative correlation (*r* = −0.250, *P* = 0.030) in **left transverse temporal cortex** that showed increase of PAC with **right** hippocampus in AD group. A linear regression model and the corresponding 95 % confidence interval is plotted in each plot for better visualisation.

## 4. DISCUSSION

This study approached Alzheimer’s disease as a disorder of disrupted communication within large-scale neural systems. Grounded in this network-based perspective, we sought to investigate whether hippocampo-cortical PAC shows measurable dysfunction in AD resting state activity. We used a principled approach for identifying genuine across-site PAC, antisymmetrized cross-bicoherence, and a pipeline that enabled reliable reconstruction of deep sources such as the hippocampus from MEG data. We found that AD affects the communication between hippocampal and cortical oscillations. We have also observed associations between these PAC alterations and cognitive ability as indexed by MMSE score in the AD group. Collectively, our findings suggest that hippocampo-cortical PAC offers a promising window into the functional connectivity disrupted by Alzheimer’s disease—and that non-invasive MEG may play an important role in uncovering cortical and deep sources connectivity. The implications of which are discussed below.

The mechanistic role of across-site PAC between hippocampal PAC neurons to ventromedial prefrontal cortex local field potential signals in control of working memory reported by Daume *et al.*^11^ and the memory deficit in AD align with our findings. Subregions of the prefrontal cortex – left frontal pole for both hippocampi and the right rostral middle frontal cortex for the right hippocampus – that exhibited significant reduced across-site PAC between the AD and control groups suggest potential hippocampal-frontal impairments in AD during the resting state. However, it is important to note that the recording site within the prefrontal cortex in Daume *et al.*^11^ was restricted to the ventromedial prefrontal cortex, leaving open the question of whether similar coupling patterns exist in other prefrontal subregions.

Among the regions exhibiting reduced coupling with the hippocampus in AD, the parahippocampal gyrus and the posterior cingulate cortex – two clusters identified in both hippocampal models – stand out as key components of the default mode network (DMN), a brain system involved in internally directed cognitive processes. Notably, the most compelling link between clinical disease and DMN disruption occurs in AD, where consistent alterations in this network have been associated with disease progression and cognitive decline.^57^ Impaired hippocampal PAC with the parahippocampal gyrus and posterior cingulate cortex in the right hemisphere may thus reflect this broader DMN disruption in AD.

The DMN comprises a set of functionally interconnected brain regions, including the ventromedial prefrontal cortex, posterior cingulate cortex, inferior parietal lobule, lateral temporal cortex, dorsal medial prefrontal cortex, and the hippocampal formation – which includes the entorhinal and surrounding parahippocampal cortex.^57^ While fMRI studies have linked DMN disruptions in AD to widespread neural, metabolic, and hemodynamic changes,^29^ MEG-derived DMN representations also show a spatial structure highly consistent with fMRI-derived maps.^58^ These oscillatory alterations within the DMN are frequency-dependent and correlate with cognitive decline.^59,60^ Furthermore, altered functional connectivity of the DMN has been observed in AD compared to MCI in eyes-closed resting-state EEG,^61^ with changes in medial temporal lobe connectivity significantly correlating with MMSE scores. Taken together, the present findings suggest that hippocampal-cortical PAC impairments within the DMN in AD may serve as indicators of disrupted network-level communication, reflecting the broader functional disintegration characteristic of disease progression.

The only region that prominently showed increased coupling in AD – in both left and right hippocampi models – is the left transverse temporal, a region associated with phonological processing. An fMRI study of AD patients on verbal short-term memory reported reduced activation of transverse temporal gyri during the encoding phase.^56^ In contrast, AD patients showed increased activation in the hippocampus, suggesting that AD patients may recruit alternative recognition mechanisms when performing a short-term memory task. However, a clear functional relation between the hippocampus and transverse temporal in resting state is missing.

An important question that arises from the results is, “Why do left and right hippocampal PAC analyses sometimes implicate the same cortical regions? Could these findings reflect merely local gamma power differences rather than hippocampal PAC?” As discussed in the Introduction section, the low signal-to-noise ratio (SNR) of gamma oscillations in resting-state MEG limits the reliable quantification of gamma activity and its group-level comparisons. Additionally, as shown in the Results section, no significant differences in gamma power were observed between groups. It is noteworthy that the study by Prabhu *et al.*^9^ proposes that PAC could be a more sensitive approach in AD versus healthy discrimination compared with the direct measurement of local gamma activity, particularly in conditions with low SNR. In conclusion, across-site PAC by providing a mechanism for the organization of activity across brain regions^10^ and a higher sensitivity in comparison with other first order measures can shed light into network vulnerability associated with AD.

Moreover, this analysis should be extended to larger, healthy cohorts to establish normative baselines and enhance statistical robustness. A systematic investigation of hippocampo-cortical PAC in healthy aging would clarify inter-individual variability and help detect deviations that signify pathological trajectories. Finally, task-based studies involving memory encoding and retrieval allow researchers to link changes in PAC more directly to behavioural impairments. Comparing resting-state and task-evoked PAC could distinguish between trait-like network deterioration and dynamic compensatory mechanisms.

### 4.1 Limitations

This exploratory study utilizes an innovative methodology involving antisymmetrized bispectrum analysis, hippocampal source estimation, and rigorous statistical testing. However, it also has limitations.

One limitation arises from the dataset provided by Kudo *et al.*^29^. The data were low-pass filtered at 55 Hz despite a sampling rate of 600 Hz, restricting the analysis to the low-gamma range. In contrast, studies such as Daume *et al.*^11^ and Wang *et al.*^10^ studies have considered gamma oscillations up to and above 100 Hz.

Additionally, the short duration of the recordings posed a challenge. To overcome this, 1-second segments with a 0.2-second shift were used for Fourier and bispectral analysis as opposed to 2-second segments that could increase the frequency resolution to 0.5 Hz. However, further testing on the random samples from Cam-CAN dataset^42^ – resting-state MEG – which includes 9-minute recordings, revealed significantly lower amounts of noise in PAC estimates when longer data segments were analysed. This suggests that longer recordings might improve the robustness of the analysis.

A further notable limitation of this study is the lack of a systematic dissociation between harmonic and non-harmonic PAC—that is, PAC arising from signals containing a periodic but non-sinusoidal component, where harmonics appear only at integer multiples of the base frequency. Giehl *et al.*^62^ found no evidence of non-harmonic within-site PAC in resting-state eyes-open MEG. They recommend bicoherence due to its high spectral resolution to distinguish between harmonic and non-harmonic PAC. Moreover, Giehl *et al.*^62^ suggest that the concurrent presence of bicoherence, amplitude-amplitude coupling (AAC), and phase-phase coupling (PPC) between the base frequency and its harmonics strongly indicates harmonic PAC. In addition, the observed across-site PAC effects should be tested against the harmonic signal, as harmonic PAC might be present in across-site PAC when there is a local harmonic together with across-site phase coupling at the harmonic base frequency.

### 4.2 Conclusion

This study demonstrated that bispectral analysis robustly detect altered patterns of cross-frequency interaction between the hippocampus and cortical regions, revealing consistent disruptions of PAC in AD patients. Specifically, reduced across-site PAC was observed between the hippocampus and several key cortical regions, including the prefrontal, parahippocampal, and posterior cingulate cortices, highlighting widespread disruptions in long-range oscillatory coupling. Notably, these effects remained significant across both hippocampi and statistical tests, indicating robust group-level differences. Replication of power alteration patterns in AD further validated the reliability of the source reconstruction and preprocessing steps, while the absence of significant group differences in gamma power supported the interpretation that gamma PAC, rather than local power confounds, reflects a network-level marker. Altogether, our findings support the view that Alzheimer’s disease impairs not only structural integrity but also the coordination of oscillatory dynamics across distributed circuits. This study adds to the growing evidence that electrophysiological markers – particularly across regional functional connectivity measures – can contribute meaningfully to our understanding and assessment of AD.

## Acknowledgements

This work was supported by the following NIH grants: R01AG088398 to KGR; R01DC017091, R01DC013979, RF1NS100440, R21DC021557, R01DC021711, R01DC019167, and P50DC019900 to S.S.N. This result is part of a project that has received funding from the European Research Council (ERC) under the European Union’s Horizon 2020 research and innovation programme (Grant agreement No. 758985).

## Consent Statement

We used a published, de-identified and publicly available dataset from (https://doi.org/10.17605/OSF.IO/PD4H9). Informed consent was obtained from all participants or their assigned surrogate decision makers. The study was approved by the Institutional Review Board of UCSF (UCSF-IRB 10–02245).

