## Supplementary materials for "Abnormal hippocampo-cortical theta-gamma phase-amplitude coupling in Alzheimer’s disease"

### Supplementary material

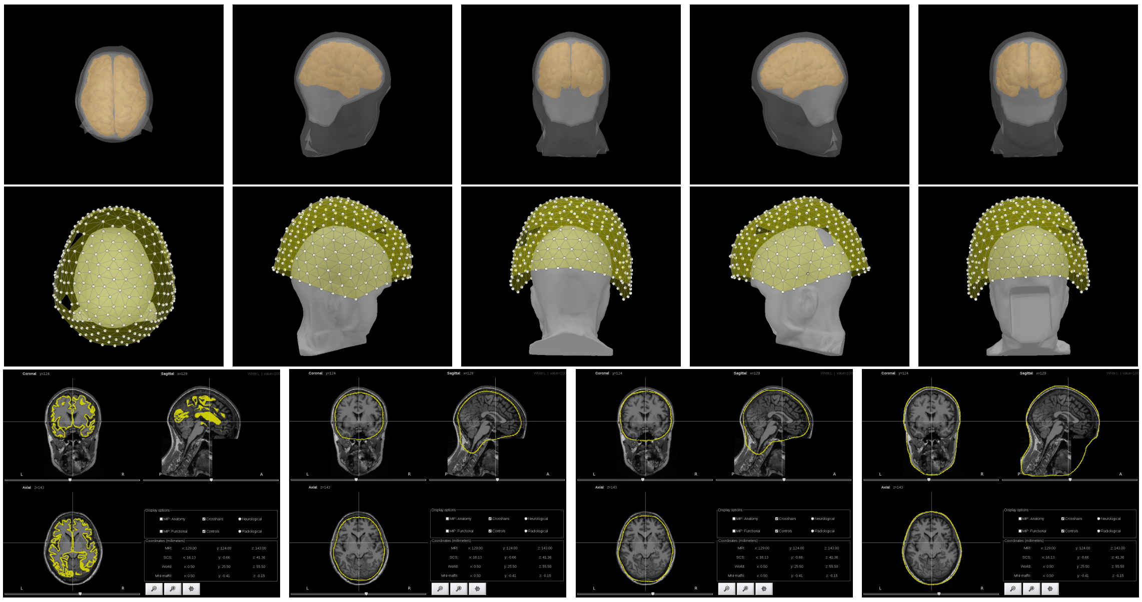

Figure 1: **Top**: BEM surfaces and cortex in different view angle, **Middle**: Overlay of sensors and reconstructed de-identified head surfaces, **Bottom**: left to right: MRI with outer cortical, inner skull (brain+CSF-skull interface), outer skull (skull-scalp interface) and head boundaries

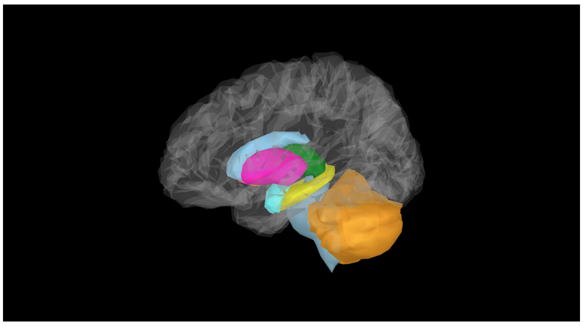

Figure 2: Subcortical volumes reconstructed by Freesurfer. The hippocampus is shown in yellow color.

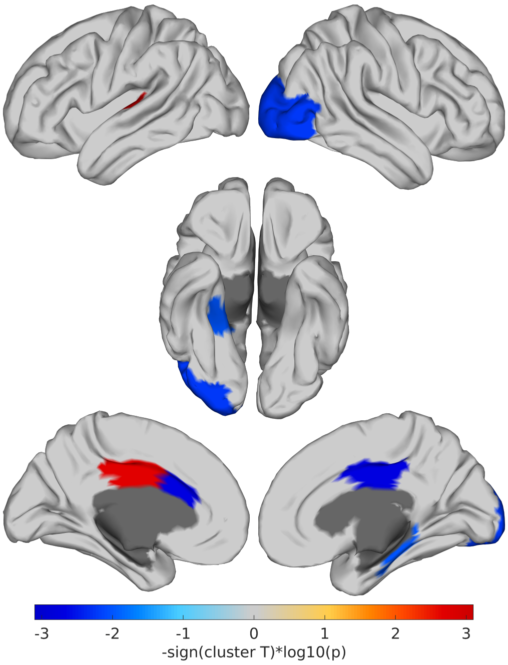


Figure 3: Group difference in **mean** across-site PAC with the **hippocampi** as the **phase** provider signal. Highlighted regions presented a significant cluster ($P < 0.01$ and cluster alpha threshold of 1%) of AD vs. control contrast identified with a **cluster based permutation test**.

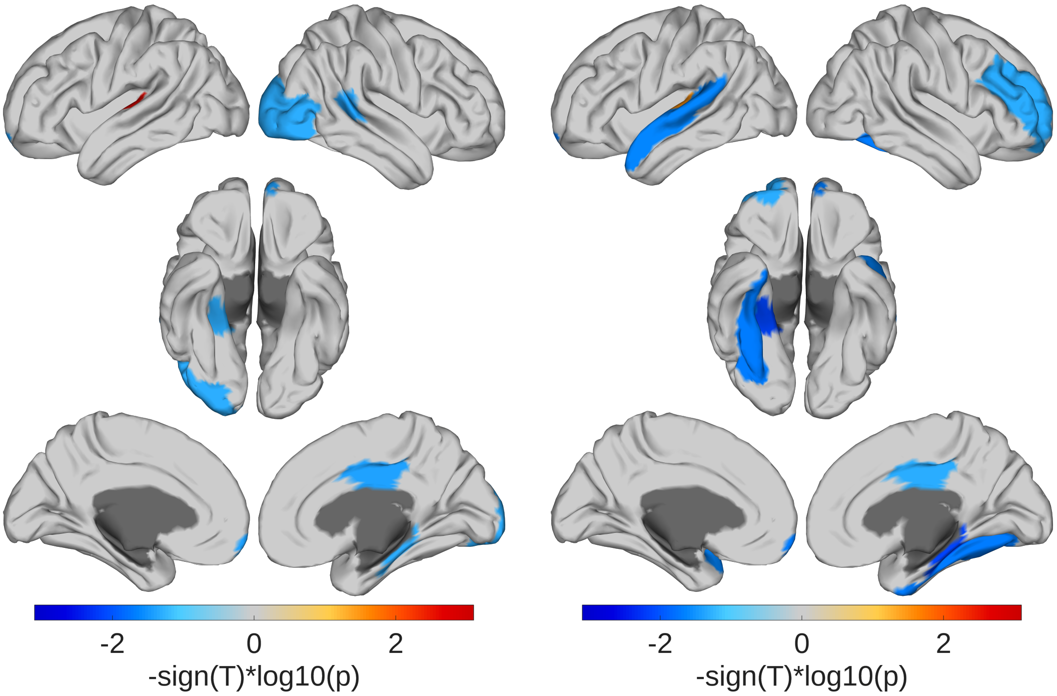


(a) **Left hippocampus** (b) **Right hippocampus**

Figure 4: Group difference in across-site PAC with the hippocampus as the **phase** provider signal. Regions identified with a **linear mixed-effects model** ($P < 0.05$ and FDR corrected)
